# A Multi-Mineral Intervention Improves Intestinal Permeability in Patients with Ulcerative Colitis: Results from a 90-Day Pilot Trial

**DOI:** 10.64898/2026.01.28.26345064

**Authors:** Muhammad N. Aslam, Danielle (Kim) Turgeon, Shannon McClintock, Ron Allen, Ananda Sen, James Varani

## Abstract

**Introduction:** Previous studies have shown that Aquamin^®^, a multi-mineral extract from red marine algae, enhances barrier integrity proteins in the human colon. These findings prompted further investigation into Aquamin^®^’s effects on gastrointestinal barrier function and permeability.

**Methods:** Subjects with mild or in remission ulcerative colitis (UC) and healthy controls were enrolled in an open-label trial and received Aquamin^®^ capsules (800 mg calcium/day) for 90 days. Intestinal permeability was evaluated before and after the 90-day intervention by urinary mannitol excretion after ingestion of a 5 g mannitol solution, with collections across several time intervals (pre-drink, 0–2 h, 2–8 h, and 8–24 h). The primary outcome was the change in mannitol excretion. Serum samples were also collected to assess liver and renal function.

**Results:** In this pilot study (NCT04855799), which included UC patients and healthy controls (n = 8 per group), baseline urine mannitol levels in the 0–2 h sample were 54% higher in UC patients compared to healthy subjects (p = 0.006). Following 90 days of Aquamin^®^ supplementation, urinary mannitol levels in UC patients decreased by 28%, 26%, and 41% at the 0–2 h, 2–8 h, and 8–24 h timepoints, respectively; the reduction at the 0–2 h interval reached statistical significance (p = 0.015). Overall, Aquamin^®^ supplementation reduced total post-intervention mannitol excretion by 29% (p = 0.024). Aquamin^®^ was well tolerated, with no serious adverse events reported. The serum metabolic panel revealed a modest but statistically significant reduction in alkaline phosphatase levels after 90 days of intervention.

**Conclusion:** These results provide preliminary evidence that Aquamin^®^ supplementation beneficially modulates gut barrier function and supports epithelial integrity in UC patients. These findings support further investigation of Aquamin^®^ as a safe and promising adjunct to current UC management strategies, with potential utility as a barrier therapy in UC.

**Summary:** Aquamin^®^ supplementation for 90 days reduced intestinal permeability in ulcerative colitis patients, as measured by urinary mannitol excretion. The intervention was well tolerated, suggesting Aquamin^®^ may be a safe, promising adjunct for enhancing gut barrier function in UC management.

## INTRODUCTION

Inflammatory bowel disease (IBD), which includes ulcerative colitis (UC) and Crohn’s disease (CD), is characterized by chronic mucosal inflammation linked to impaired intestinal barrier function and increased permeability, or “leaky gut” [1–3]. Normally, the epithelial barrier maintains selective absorption and defends against tissue influx of luminal pathogens, antigens, and toxins. In IBD, this barrier breaks down, allowing translocation of microbes and antigens into the submucosa, to fuel immune activation and ongoing inflammation. Notably, barrier defects may precede and perpetuate mucosal inflammation, acting as both cause and consequence of disease [4–8]. In UC, diffuse colonic inflammation and superficial ulcers correspond with barrier dysfunction across disease severities. Even patients in remission can have persistent symptoms associated with ongoing permeability defects, which correlate with relapse risk and response to therapy [1–3]. Thus, assessment of intestinal permeability is increasingly used for disease monitoring and management in UC.

Given this central pathogenic role, interest has grown in therapies aimed at restoring epithelial integrity and improving barrier function in UC. Numerous micronutrients, traditional remedies and natural product supplements have been evaluated for their ability to improve permeability control and support gut barrier health [2,9]. Despite encouraging results in experimental models and preliminary human studies, however no intervention has yet to demonstrate sufficient efficacy needed to justify widespread adoption [2].

For the past several years, our laboratory has investigated the potential of a calcium-rich, magnesium-rich, multi-mineral product known as Aquamin^®^ to support gastrointestinal (GI) health. In long-term (15-18 month) interventional studies in mice, inclusion of Aquamin^®^ in the diet suppressed polyp formation throughout the GI tract [10,11] as well as liver tumor formation [12]. A reduction in GI and systemic inflammation was seen in conjunction with these health benefits [10–14]. Consistent with evidence of anti-inflammatory potential, Aquamin^®^ has also demonstrated the ability to suppress colitis development in the IL-10^-/-^ mouse model [15].

How the multi-mineral product promotes GI health is not fully understood. Studies using human colonic tissue in organoid culture (derived from both healthy individuals and from UC patients) have demonstrated increased elaboration of multiple proteins that contribute to barrier structure/function [16–20]. An increase in trans-epithelial electrical resistance (TEER) and enhanced organoid cohesion accompanied these protein changes [18–20]. Barrier proteins remained upregulated with Aquamin^®^ even when the colon organoids were challenged with a mix of LPS and three pro-inflammatory cytokines [20,21]. Most importantly, many of the same barrier protein changes observed in human colon tissue-derived organoid cultures were also observed in colon biopsies obtained from healthy subjects immediately following daily ingestion of Aquamin^®^ over a 90-day period [22].

Based on the findings presented above, we carried out a 180-day interventional trial in patients with UC in remission or with mild disease (clinicaltrials.gov ID: NCT03869905) [23]. The goal of the trial was to determine if disease-related and mechanistic biomarkers could be improved in this population with Aquamin^®^. Over the 180-day treatment period, we saw reductions in several disease-related biomarkers that are typically elevated in UC. Among these were C-reactive protein (CRP), total and intestinal alkaline phosphatase (ALP), and fecal-calprotectin. Histological evidence of reduced colonic inflammation was also seen. These changes were not observed in patients receiving placebo. Colon tissue biopsies obtained at the end of the interventional period also demonstrated increased expression of several gut barrier proteins compared to pre-intervention levels in the Aquamin^®^-treated cohort. These changes were not observed with placebo. The results from the interventional study allow us to suggest that Aquamin^®^ could serve as part of a maintenance regimen for individuals with mild UC or in remission [23].

The findings presented in our interventional trial along with the preclinical findings also allow us to hypothesize improvement in gastrointestinal barrier function as (at least) part of Aquamin^®^’s mechanism of action. To date, however, there is no direct evidence demonstrating improved barrier function *in vivo*. As a way to address this issue, mannitol recovery in the urine over a 24-hour period following ingestion of the probe was assessed in a cohort of subjects (8 healthy individuals and 8 subjects with mild UC or UC in remission) before and after 90 days of Aquamin^®^ treatment under the same conditions used in the recent clinical study described above [23]. The results of this study are described here.

## MATERIALS AND METHODS

### Intervention (Aquamin^®^)

This open-label interventional study utilized Aquamin^®^ following its designation as an Investigational New Drug (IND# 141600) by the U.S. Food and Drug Administration (FDA). Aquamin^®^ is a product rich in calcium, magnesium, and numerous trace minerals, and is sourced from calcified fronds of marine red algae belonging to the Lithothamnion genus [24]. The formulation contains calcium and magnesium in approximately a 12:1 ratio, along with measurable amounts of seventy-two additional trace elements. Marketed as a dietary supplement (GRAS 000028; Marigot Ltd., Cork, Ireland), Aquamin^®^ is incorporated into foods and supplements available in Europe, Asia, Australia, and North America. Aquamin^®^ is available in multiple different preparations; for this trial, a single batch of Aquamin-TG^®^ (Food Grade) was encapsulated in hydroxypropyl methylcellulose (HPMC). Each capsule contained 600 mg of Aquamin^®^ standardized to deliver 200 mg elemental calcium. The University of Michigan Research Pharmacy dispensed a 90-day supply of Aquamin^®^ to participants. Each participant took four capsules per day, two in the morning and two in the evening, to achieve a total daily intake of 800 mg calcium, administered in addition to their ongoing UC maintenance therapy. The full elemental profile and daily intake amounts (of Aquamin-TG^®^) are presented in Supplementary Table 1. The mineral content of Aquamin-TG^®^ was verified by independent analysis (Advanced Laboratories, Salt Lake City, Utah) using Inductively Coupled Plasma Optical Emission Spectrometry; this intervention has been utilized in previous clinical research [22,23,25].

### Study design

This single-site study was a pilot-phase, open-label interventional trial that included participants with UC, either in remission or with mild disease, as well as healthy individuals. The primary objective was to determine whether daily Aquamin^®^ supplementation for 90 days could improve gastrointestinal barrier function, as assessed by mannitol recovery in the urine of healthy subjects and those with UC.

### Regulatory oversight

This investigator-initiated interventional study was conducted under FDA approval, with JV acting as the study sponsor. The Institutional Review Board at the University of Michigan Medical School (IRBMED, HUM#00156676) provided regulatory oversight. This study was conducted as a substudy of an ongoing trial (ClinicalTrials.gov ID: NCT03869905) and under the same HUM#00156676 [23]. The Michigan Institute for Clinical and Health Research (MICHR) supported site monitoring, data collection, and data analysis using the REDCap (Research Electronic Data Capture) platform. A dedicated Data and Safety Monitoring Committee (DSMC) conducted monthly reviews to ensure participant safety and maintenance of data integrity. The trial was registered as an interventional clinical study on ClinicalTrials.gov (NCT04855799) in April 2021, with the first participant enrolled in November 2021 and the last completing the study in March 2024. All participants provided written informed consent before enrollment and prior to any intervention. The study was conducted in accordance with established ethical standards, including the Declaration of Helsinki, CIOMS International Ethical Guidelines for Biomedical Research Involving Human Subjects, ICH Good Clinical Practice, the Belmont Report, and the U.S. Common Rule.

### Study participants

Study participants were recruited through the Michigan Medicine IBD clinics, the UMHealthResearch web portal, and by posting flyers in the hospital. Figure 1 presents a CONSORT flow diagram detailing subject enrollment, randomization, and intervention allocation. The initial intent was to include 20 evaluable subjects (10 per arm) in this pilot-phase study. Although there was no prior clinical data to support this number, a previous trial with healthy subjects demonstrated that statistically significant differences in proteomic findings could be observed with as few as ten subjects per arm [22]. Based on the expected dropout and screen failure rates, a total of 31 subjects were screened, and 29 subjects were enrolled in this interventional trial. Two subjects failed screening. Seven subjects withdrew, and the remaining 22 were allocated to the intervention. Of those who started on treatment, 21 completed the study, while one discontinued the treatment. Five subjects were considered inevaluable at study completion due to multiple missing urine samples, combining urine samples from two timepoints, or intake of fructose-containing beverages during the collection window. As a result, only sixteen subjects were fully evaluable after 90 days of intervention. This included eight subjects in each group (healthy individuals and subjects with UC) who received Aquamin^®^ for the entire 90-day period.

**Figure 1.**
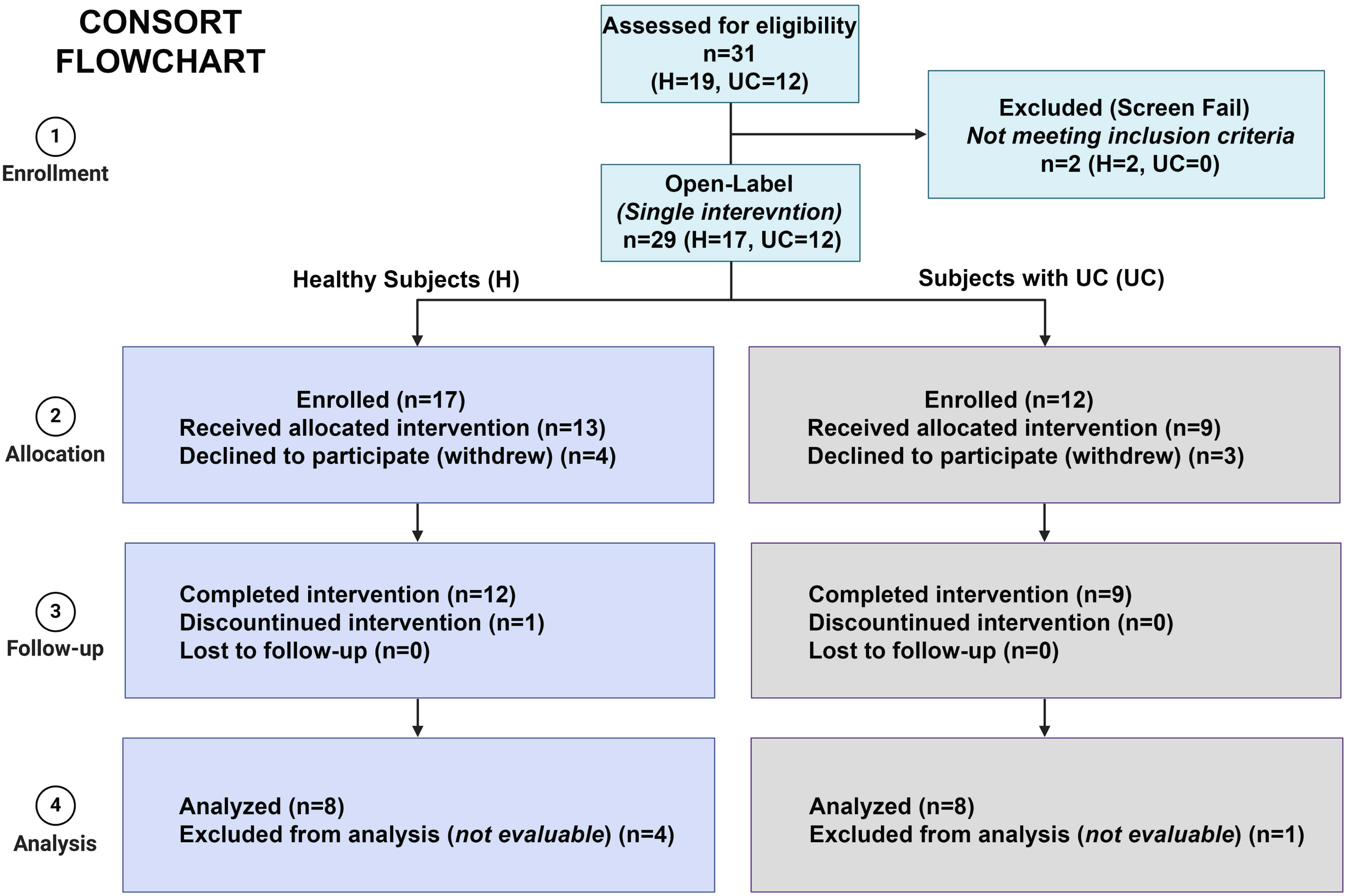
CONSORT flow diagram. Participant screening, enrollment, allocation, follow-up, and analysis for the 90-day open-label, single-intervention study in healthy subjects (H) and subjects with ulcerative colitis (UC). Final evaluable sample sizes, with withdrawals, discontinuations, and exclusions as not evaluable are shown.

Participants were males or non-pregnant females in generally good health, or individuals with a confirmed diagnosis of UC. Eligible individuals with UC were either in remission or had mild disease at enrollment, as determined by the study gastroenterologist (DKT), with stable maintenance therapy and an Inflammatory Bowel Disease Questionnaire (IBDQ) score of 170 or above. The IBDQ [26] assesses health-related quality of life for UC. The IBDQ evaluates bowel, systemic, emotional, and social functions, with scores of 170 or above often indicating remission. Supplementary Table 2 presents the IBDQ scores at baseline, reflecting disease status prior to the start of the intervention.

Participants were excluded from both the healthy and UC cohorts if they were pregnant, lactating, or of childbearing potential and unwilling to use acceptable birth control throughout the study period. Individuals were also excluded if they were participating in any other interventional trial involving an investigational drug, or if they were deemed unlikely to cooperate with or comply with study procedures. Subjects with a history or diagnosis of CD, other IBDs, functional gastrointestinal disorders (including irritable bowel syndrome [IBS]), active UC within three months prior to enrollment (except for mild cases), gastrointestinal bleeding disorders (including those from gastric or duodenal ulcers or gastrin-secreting tumors), peptic ulcer disease with bleeding in the preceding three months, any gastrointestinal or colonic malignancy, kidney disease (including kidney stones or hypercalcemia), coagulopathies, hereditary hemorrhagic disorders, or neurologic disease were not eligible for participation.

Furthermore, individuals were excluded if, within 30 days prior to study entry, they had taken or were unwilling to forgo for 30 days prior to enrollment any supplements containing calcium, vitamin D (including multivitamins with these nutrients), magnesium, or fiber, non-steroidal anti-inflammatory drugs (NSAIDs; except for occasional pain management or low-dose aspirin for cardiovascular disease prevention), corticosteroids, or antibiotics.

### Study protocol

Figure 2 provides an overview of the study design. Each participant attended three study visits: (i) screening and enrollment, (ii) baseline (Day 0), and (iii) the final visit (Day 90). At the screening visit, potential participants received a full explanation of the study, after which they were administered the NIH Diet History Questionnaire III (DHQ III) [epi.grants.cancer.gov/dhq3], a comprehensive food frequency questionnaire incorporating portion size and dietary supplement intake that was used to assess baseline calcium and selected mineral intake over the previous 12 months [22,23,25]. Participants were also queried regarding their consumption of dietary supplements, antibiotics, steroids and NSAIDs. A medical and medication history was obtained, followed by a brief physical examination. Written informed consent was obtained from eligible individuals prior to any study-related procedures. At both Visit 2 (baseline, Day 0) and Visit 3 (final, Day 90), participants provided serum samples (by venous blood draw), as well as 24-hour urine samples collected. The IBDQ was completed at both visits to assess disease-related quality of life. Monthly monitoring occurred on Day 30 and Day 60, during which toxicity, any adverse event and adherence were assessed.

**Figure 2.**
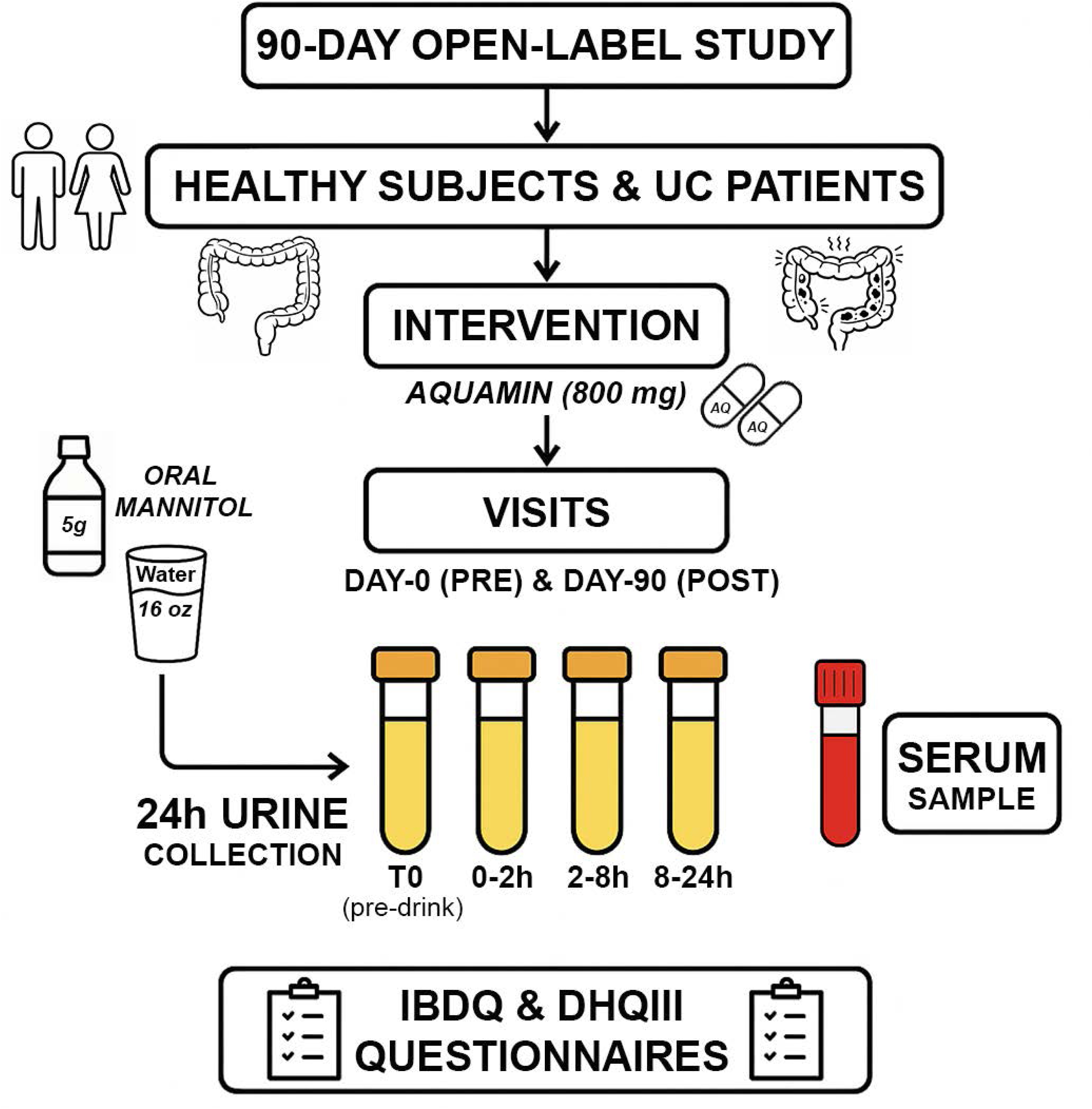
Study design and sample-collection schematic. Overview of the 90-day open-label study in healthy subjects and UC patients receiving Aquamin^®^ (800 mg of calcium). Study visits occurred at Day 0 (pre) and Day 90 (post). Gastrointestinal permeability was assessed using an oral mannitol challenge (5 g in 16 oz water) followed by 24-hour urine collection partitioned into T0 (pre-drink), 0–2 h, 2–8 h, and 8–24 h fractions; a serum sample and IBDQ questionnaires were also collected. DHQIII survey was also offered to assess dietary calcium intake for the last 12 months.

### 24-hour urine test for mannitol recovery (GI permeability assessment)

Intestinal permeability was evaluated by measuring urinary recovery of D-mannitol, a non-metabolized sugar that served as the probe. The University of Michigan Research Pharmacy dispensed D-mannitol (5 g of water-soluble crystalline powder; MEDISCA Inc., product no. 0599; chemical name: 1,2,3,4,5,6-hexanehexol).

Urinary recovery of mannitol was measured at both Visit 2 (baseline, Day 0) and Visit 3 (final, Day 90) by performing a 24-hour urine collection. On the morning of each visit, participants collected a pre-drink (pre-mannitol) urine sample following an overnight fast (of 6-8 hours), placing the sample in a designated container kept on ice or under refrigeration. After providing the pre-drink sample, participants consumed the test solution containing 5 g D-mannitol dissolved in 16 oz of water, ingested over a period of 15–20 minutes, immediately before the start of the 24-hour urine collection. They were allowed to take one additional cup (16 oz) of water before or during that period. Following administration of the test solution, participants abstained from both food and beverage intake (other than water) for the first eight hours of the subsequent 24-hour urine collection period. Participants were allowed to drink two additional cups of water (16 oz each) after the first two hours (during the next 6 hours) of urine collection. All urine excreted during the 24-hour period was collected in separate containers according to the following schedule: pre-mannitol urine in container #1, the first two hours in container #2, hours 2–8 in container #3, and hours 8–24 in container #4. All containers were kept chilled throughout the collection period. At the end of the initial eight-hour window, participants resumed their regular food and water intake, avoiding only fructose-containing foods and documenting all fluid consumption to account for variability.

Participants were instructed to maintain their usual diet and fluid intake the day prior to testing, with the exception of strict avoidance of specific foods and beverages for at least 24 hours before the test. These restricted items included fruits, fruit juices, jams, jellies, foods and beverages sweetened with high-fructose corn syrup, artificial sweeteners, alcoholic drinks, dairy products, dietetic chocolate, honey, mushrooms, legumes (including beans and peanuts), celery, and chewing gum. Additionally, all marijuana-related products and mannitol-containing foods or supplements were prohibited for at least 48 hours prior to and during testing.

Upon returning to the study center, participants submitted their complete 24-hour urine collections. The total volume of each sample was measured, and aliquots were prepared for storage. All samples were promptly frozen at –80°C. Mannitol recovery and intestinal permeability were assessed at the end of the study by quantifying urinary mannitol concentration using the ELISA kit (ChromaDazzle D-Mannitol Assay Kit by AssayGenie; Cat# BA0133).

### Serum Biomarkers

A comprehensive metabolic panel was performed to assess serum biomarkers, including total albumin, bilirubin, aspartate aminotransferase (AST), alanine aminotransferase (ALT), and ALP, along with additional analytes relevant to liver and kidney function. For participants with UC, CRP levels were measured concurrently with the metabolic panel. All serum analyses were conducted by the Michigan Medicine laboratory, which generated individual reports for each participant in accordance with established standard operating procedures. The comprehensive metabolic panel was evaluated primarily to monitor safety, although CRP and ALP measurements were also clinically relevant for routine UC management.

### Statistical evaluation

This pilot study was designed to assess feasibility and tolerability of Aquamin^®^, as well as to evaluate biomarkers related to gastrointestinal permeability. We anticipated that the study would reveal trends useful for estimating effect sizes and guiding the formulation of research hypotheses for subsequent, fully powered, large-scale trials. It was hypothesized that Aquamin^®^ would exert a positive effect on mannitol recovery, with the magnitude of the effect varying among participants.

Based on the study design, pre- and post-intervention values for various serum analytes included in the metabolic panel, as well as CRP and urinary mannitol levels (measured by ELISA), were obtained from each subject. Group means and standard deviations were calculated for each endpoint, and within each cohort, pre-versus post-intervention data were analyzed using paired t-tests (at the 95% confidence level). Subsequently, an independent-samples t-test was conducted to compare the pre-post change score between the two cohorts. To account for inflated Type I error due to testing multiple endpoints, Tukey’s multiple comparisons test was used to determine significance. A p-value <0.05 was considered significant. Statistical analyses were conducted using GraphPad Prism v10.2. Given the small sample size, analyses were not adjusted for baseline sociodemographic variables, including gender, or for initial dietary calcium intake levels.

## RESULTS

### Participant demographics and baseline characteristics

Out of 31 screened subjects, sixteen subjects (eight healthy individuals and eight with UC, either in remission or with mild disease) completed the study and were considered evaluable. Supplementary Table 3 (and Figure 1) presents demographic profiles (including ethnicity) of subjects who were assessed for eligibility and enrolled in the 90-day interventional study. Of these 31 subjects, sixteen were female and fifteen were male, with comparable numbers of males and females enrolled in both the healthy and UC cohorts. The mean age of healthy subjects was 31.7 (± 12.0) years, while the mean age for those with UC was 41.9 (± 12.7) years at the start of the study. Body mass index (BMI) remained unchanged in both cohorts over the

### 90-day study period compared to baseline

Supplementary Table 4 presents average daily intakes for eight minerals, estimated from participant dietary reports over the previous 12 months using the NIH DHQ III, compared to recommended guidelines. Both healthy subjects and those with UC exhibited similar intake patterns for most minerals, generally meeting or exceeding suggested targets for calcium, copper, zinc, iron, and selenium, but falling short of recommendations for potassium and vitamin D. Magnesium intake was below recommended levels in healthy subjects but adequate among UC subjects. Notably, while vitamin D intake from food and supplements (estimated by DHQIII) remained substantially below recommended levels in both groups, the DHQIII assessment did not include questions related to sun exposure (see Supplementary Table 4).

### Participant Safety

For safety assessment, subjects reported side effects experienced during the study, and serum samples were collected to evaluate liver and kidney function.

Adverse events (AE) were reported in six healthy subjects (out of 13) and six UC subjects (out of 9), with a total of 20 and 10 events, respectively (Supplementary Table 5). No serious AEs occurred during the study. Most events were considered unrelated to the study except for gastrointestinal symptoms which included flatulence, abdominal discomfort, and constipation. These results align with our previous findings in both healthy individuals [22,25] and subjects with UC [23], indicating that Aquamin^®^ is safe to use and unlikely to pose safety or tolerability concerns. Two adverse events were also reported following mannitol administration, including diarrhea (loose stool) in one UC subject (Supplementary Table 5).

A metabolic panel was performed on serum samples from each participant at study initiation and after the final dose of Aquamin^®^ on Day-90. Results are presented in Table 1. Serum metabolic panel values for both healthy participants and those with UC remained within normal reference ranges from baseline to post-intervention, with no clinically significant changes observed over the 90-day study period. Mean values for major safety markers—including liver enzymes (AST, ALT, ALP), kidney function indicators (BUN, creatinine), and electrolytes—were stable, suggesting no adverse impact of the intervention. Overall, no trends indicating treatment-related hepatic or renal toxicity were observed in either cohort, consistent with findings from our previous trials [22,23,25]. Also, in line with what was reported previously [23], there was a small but consistent decrease in serum ALP levels with Aquamin^®^ over the course of the 90-day treatment period: 11.9% in healthy subjects and 3.3% in UC subjects.

**Table 1.**
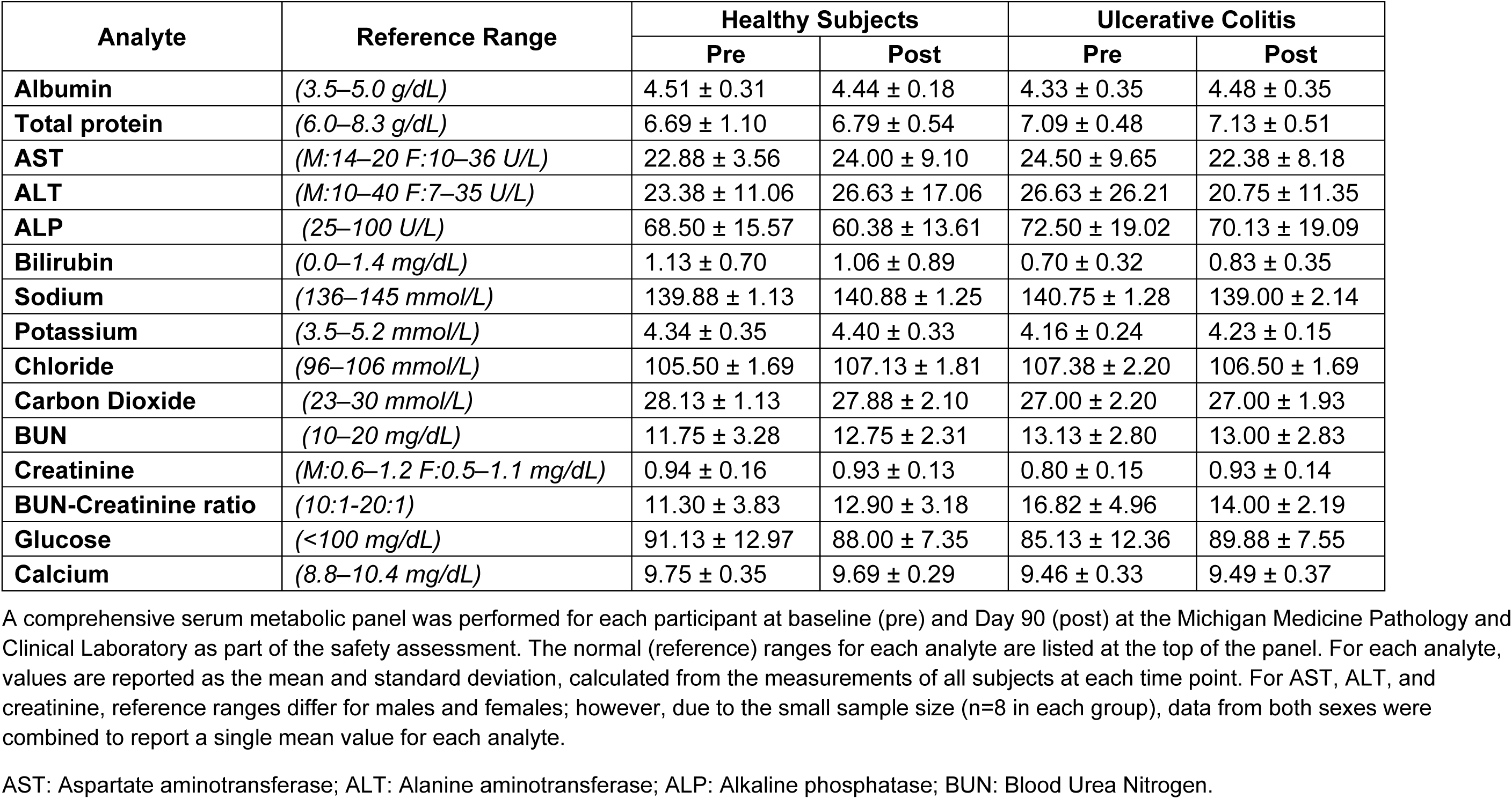
Serum chemistry (metabolic panel)

| Analyte | Reference Range | Healthy Subjects |  | Ulcerative Colitis |  |
| --- | --- | --- | --- | --- | --- |
|  |  | Pre | Post | Pre | Post |
| **Albumin** | (3.5–5.0 g/dL) | 4.51 ± 0.31 | 4.44 ± 0.18 | 4.33 ± 0.35 | 4.48 ± 0.35 |
| **Total protein** | (6.0–8.3 g/dL) | 6.69 ± 1.10 | 6.79 ± 0.54 | 7.09 ± 0.48 | 7.13 ± 0.51 |
| **AST** | (M:14–20 F:10–36 U/L) | 22.88 ± 3.56 | 24.00 ± 9.10 | 24.50 ± 9.65 | 22.38 ± 8.18 |
| **ALT** | (M:10–40 F:7–35 U/L) | 23.38 ± 11.06 | 26.63 ± 17.06 | 26.63 ± 26.21 | 20.75 ± 11.35 |
| **ALP** | (25–100 U/L) | 68.50 ± 15.57 | 60.38 ± 13.61 | 72.50 ± 19.02 | 70.13 ± 19.09 |
| **Bilirubin** | (0.0–1.4 mg/dL) | 1.13 ± 0.70 | 1.06 ± 0.89 | 0.70 ± 0.32 | 0.83 ± 0.35 |
| **Sodium** | (136–145 mmol/L) | 139.88 ± 1.13 | 140.88 ± 1.25 | 140.75 ± 1.28 | 139.00 ± 2.14 |
| **Potassium** | (3.5–5.2 mmol/L) | 4.34 ± 0.35 | 4.40 ± 0.33 | 4.16 ± 0.24 | 4.23 ± 0.15 |
| **Chloride** | (96–106 mmol/L) | 105.50 ± 1.69 | 107.13 ± 1.81 | 107.38 ± 2.20 | 106.50 ± 1.69 |
| **Carbon Dioxide** | (23–30 mmol/L) | 28.13 ± 1.13 | 27.88 ± 2.10 | 27.00 ± 2.20 | 27.00 ± 1.93 |
| **BUN** | (10–20 mg/dL) | 11.75 ± 3.28 | 12.75 ± 2.31 | 13.13 ± 2.80 | 13.00 ± 2.83 |
| **Creatinine** | (M:0.6–1.2 F:0.5–1.1 mg/dL) | 0.94 ± 0.16 | 0.93 ± 0.13 | 0.80 ± 0.15 | 0.93 ± 0.14 |
| **BUN-Creatinine ratio** | (10:1-20:1) | 11.30 ± 3.83 | 12.90 ± 3.18 | 16.82 ± 4.96 | 14.00 ± 2.19 |
| **Glucose** | (<100 mg/dL) | 91.13 ± 12.97 | 88.00 ± 7.35 | 85.13 ± 12.36 | 89.88 ± 7.55 |
| **Calcium** | (8.8–10.4 mg/dL) | 9.75 ± 0.35 | 9.69 ± 0.29 | 9.46 ± 0.33 | 9.49 ± 0.37 |

A comprehensive serum metabolic panel was performed for each participant at baseline (pre) and Day 90 (post) at the Michigan Medicine Pathology and Clinical Laboratory as part of the safety assessment. The normal (reference) ranges for each analyte are listed at the top of the panel. For each analyte, values are reported as the mean and standard deviation, calculated from the measurements of all subjects at each time point. For AST, ALT, and creatinine, reference ranges differ for males and females; however, due to the small sample size (n=8 in each group), data from both sexes were combined to report a single mean value for each analyte.
AST: Aspartate aminotransferase; ALT: Alanine aminotransferase; ALP: Alkaline phosphatase; BUN: Blood Urea Nitrogen.

When pre- and post-intervention differences in serum ALP from all 16 participants (all on Aquamin^®^) were analyzed together using a paired t-test, the overall decrease (7.4%) reached statistical significance (p = 0.026; Figure 3).

**Figure 3.**
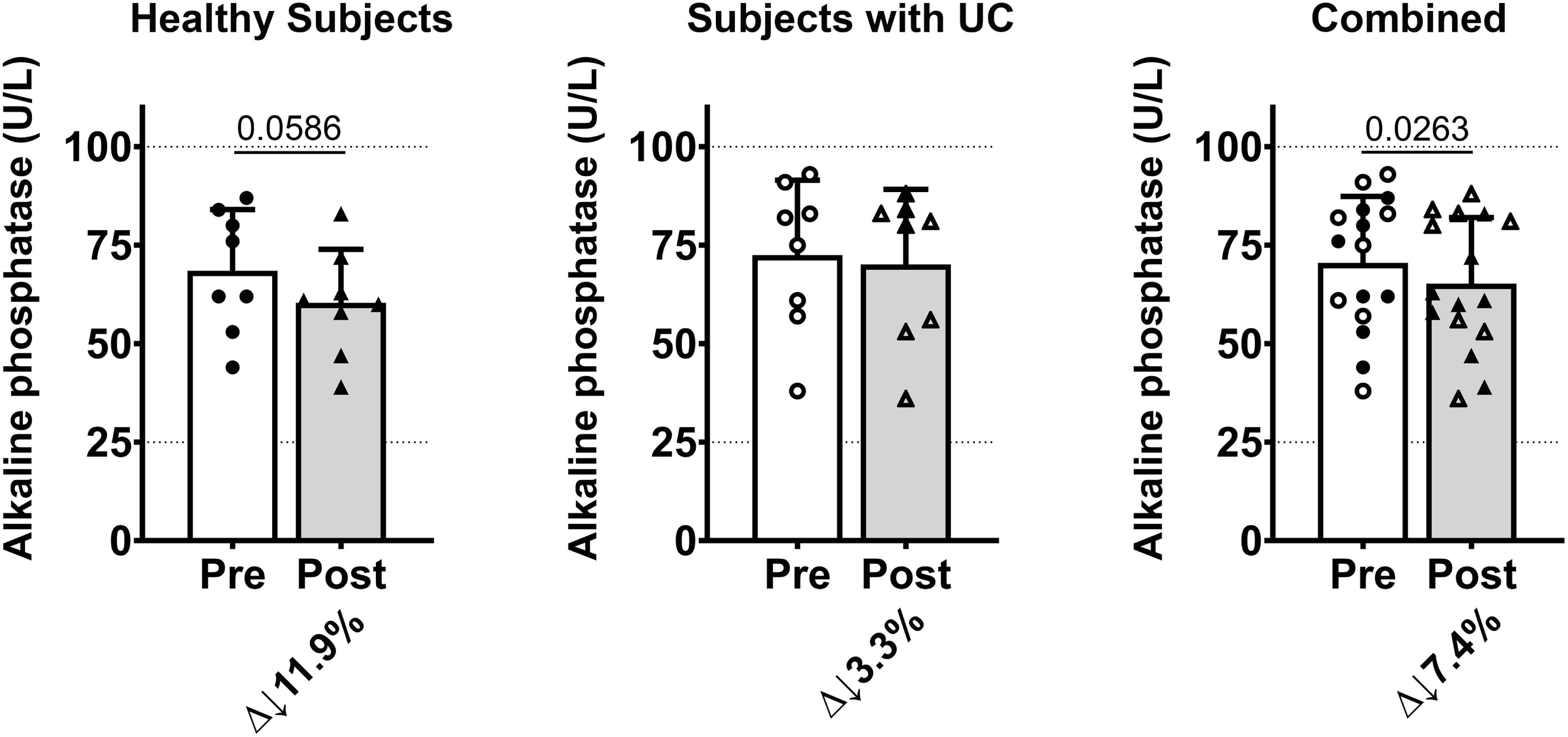
Serum alkaline phosphatase pre-vs post-intervention. Serum alkaline phosphatase (ALP; U/L) was measured at baseline (Pre) and after 90 days (Post) in healthy subjects, UC subjects, and the combined analysis. Bars show the group mean with standard deviation error bars, and points indicate individual participant values. Pre–post percent change (Δ) is shown below each panel, and two-tailed paired t-test P values for the pre–post comparison are shown above.

Serum CRP was assessed only in subjects with UC (in remission or having mild disease). At study completion, CRP values were 0.39 ± 0.13, compared to baseline values of 0.40 ± 0.19, representing a 3% decrease. This reduction is similar to that observed in our previous study of UC patients (in remission or with mild disease) after 90 days of Aquamin^®^ intervention [23]. Supplementary Table 2 presents the IBDQ scores, before and after 90 days of intervention. Average group scores changed little between Day 0 and Day 90. Among UC participants who received Aquamin for 90 days, mean scores on the full questionnaire increased from 191.6 ± 15.3 to 194.3 ± 20.5, a modest trend consistent with our prior study involving UC subjects in remission or with mild disease [23].

### Gastrointestinal permeability assessment

To assess *in vivo* gastrointestinal permeability, we administered an oral mannitol dose (5 g) and measured urinary mannitol levels via ELISA at multiple timepoints. Data are presented in Figures 4 and 5.

**Figure 4.**
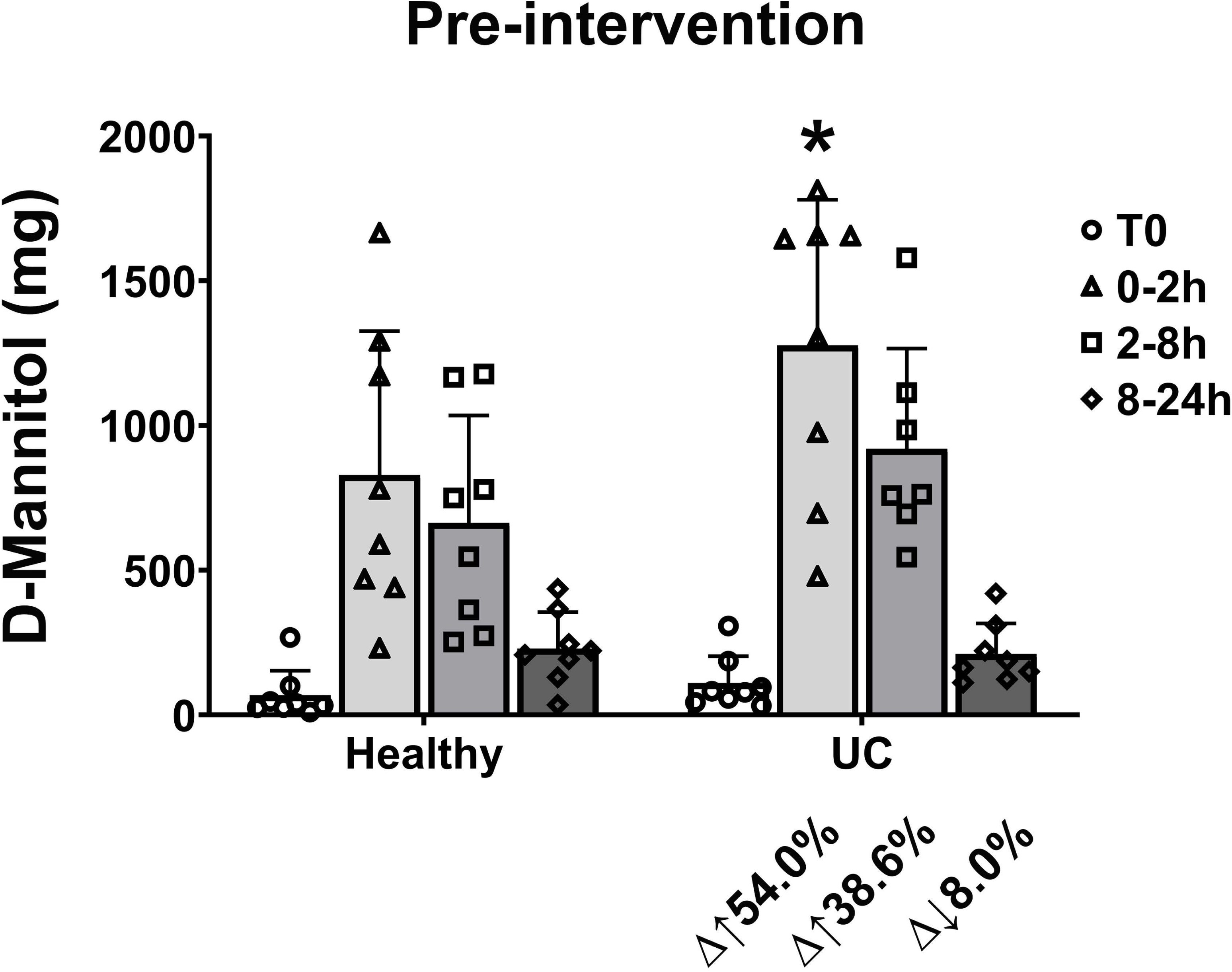
Baseline urinary D-mannitol excretion in healthy subjects and UC patients. Pre-intervention urinary D-mannitol (mg) recovered in sequential urine collections (T0, 0–2 h, 2–8 h, 8–24 h) following an oral mannitol challenge is shown for healthy subjects versus UC subjects. Bars show the group mean with standard deviation error bars; symbols denote individual participant values. Percent difference (Δ) for UC relative to the corresponding healthy interval is shown below the UC bars. * indicates significance with P<0.05 for the UC vs healthy mannitol excretion comparison at 0–2 h.

**Figure 5.**
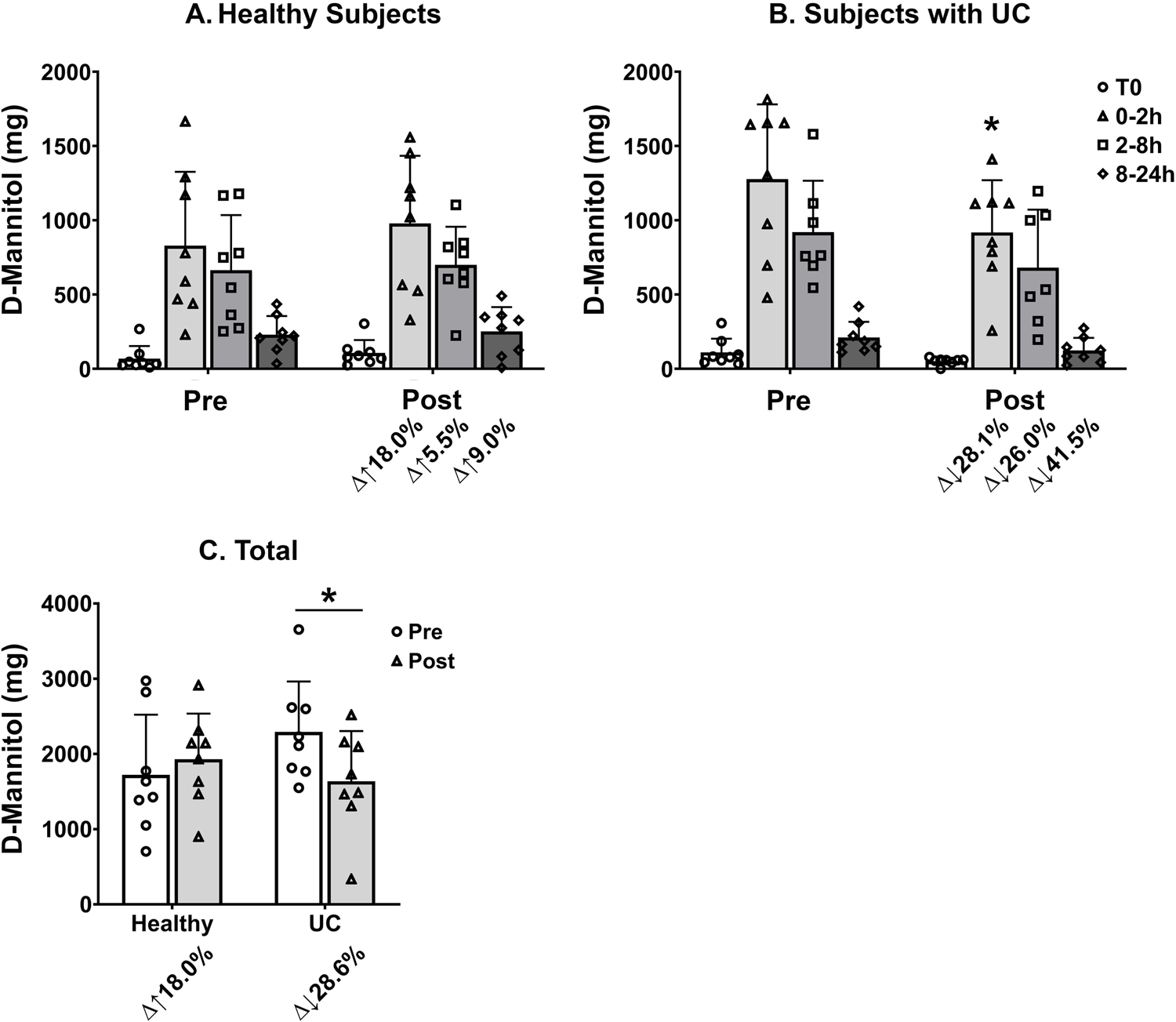
Effect of Aquamin^®^ intervention on urinary D-mannitol excretion. (A) Healthy subjects and (B) UC subjects: urinary D-mannitol (mg) in each collection interval (T0, 0–2 h, 2–8 h, and 8–24 h) at baseline (Pre) and after 90 days (Post). (C) Total 24-hour D-mannitol excretion (mg) pre- and post-intervention for each group. Bars show group mean with standard deviation error bars; symbols indicate individual participant values. Pre-post percent change (Δ) from baseline is shown; * indicates significance with P<0.05 for the indicated pre–post comparisons (in B: UC subjects, pre-post difference is significant at 0-2 h and in C: for UC subjects, total 24-h pre-post difference is significant).

Figure 4 compares pre-intervention (prior to Aquamin^®^ treatment) urinary mannitol excretion in healthy controls and subjects with UC across defined intervals – i.e., at baseline (prior to mannitol consumption and at 0-2 hours, 2-8 hours and 8-24 hours post-mannitol consumption). Baseline (T0; pre-mannitol drink) levels were minimal in both groups. In healthy participants, mannitol recovery peaked in the 0–2-hour interval (mean 829 mg), was moderate at 2–8 hours and declined further by 8–24 hours. UC subjects demonstrated significantly greater mannitol excretion at 0–2 hours (mean 1277 mg, p= 0.0064 compared to healthy counterparts at 0–2 hours), indicative of increased early intestinal permeability. Elevated excretion persisted, though at reduced levels, in subsequent intervals. Percent change analysis in UC subjects showed a 54.0% increase at 0–2 hours and a 39% increase at 2–8 hours compared to healthy subjects at the same timepoints. These results reveal a more substantial and more rapid urinary mannitol recovery in the UC cohort, most evident in the first 2 hours post-administration.

Figure 5 compares urinary mannitol excretion in healthy subjects and those with UC before and after 90 days of intervention with Aquamin^®^. In healthy subjects (Figure 5A), no significant differences in urinary mannitol excretion were observed between pre- and post-intervention across all collection intervals. Post-intervention mannitol levels were slightly increased. In UC subjects (Figure 5B), post-intervention mannitol excretion during the 0–2-hour interval was significantly reduced compared to pre-intervention values (p= 0.015), with an overall percent decrease of 28%. Mean urinary mannitol levels decreased from 1277 mg at pre-intervention to 917 mg at post-intervention. Mannitol levels continued to decline in post-intervention samples across the next two collection timepoints. Figure 5C provides a summary of the total urinary mannitol values in pre- and post-treatment specimens from the two participant groups. The combined analysis (Figure 5C) confirms a significant reduction (p= 0.0236) in total urinary mannitol recovery post-intervention for UC. That is, an overall reduction of 29% (from approximately 2300 mg at baseline to 1637 mg at day-90) was observed in the UC participant group. In healthy subjects, total mannitol recovery was virtually unchanged at day-90 as compared to pre-Aquamin^®^ intervention.

## DISCUSSION

A breakdown in the permeability barrier of the gastrointestinal tract is a feature of UC [1–3]. While bowel inflammation is recognized as a cause of barrier dysfunction, increased permeability has been observed in some UC patients even in the absence of overt inflammation [27,28]. Increased permeability has also been shown to predict disease recurrence among UC patients in remission [29] and has been correlated with subsequent disease development in first-degree relatives of UC sufferers [30]. Thus, barrier breakdown may promote inflammation in addition to being its result. Regardless of whether barrier dysfunction triggers inflammation or is a consequence of it, improving barrier integrity is likely to be beneficial. Numerous dietary supplements have been purported to reduce “leaky gut” [2,9], but to date, none has proven sufficiently effective to warrant widespread recommended use. Here we present evidence to demonstrate that daily consumption of a multi-mineral product over a 90-day period has the potential to improve barrier function in patients with mild UC or those in remission. Results from our previous organoid culture studies [16–21] and findings from two interventional trials [22,23] demonstrated upregulation of multiple proteins involved in colonic barrier integrity.

Building on these considerations, our evaluation of gut permeability in participants with mild UC or those in remission, compared with healthy subjects, demonstrates a characteristic difference at baseline. Participants with UC had greater early uptake and transfer of mannitol from the gut into the bloodstream (reflected by higher 0–2-hour urinary recovery), consistent with increased intestinal permeability (a “leakier” barrier) compared to healthy controls (Figure 4). In participants with UC, the post-intervention decreases in the 0–2-hour and total (24-hour) mannitol recovery indicate (Figure 5) that Aquamin^®^ intervention was associated with reduced permeability and improved barrier function. In contrast, the lack of change in healthy subjects suggests no material effect when baseline permeability is already normal.

Gastrointestinal permeability refers to the transport of substances across the gut wall through both transcellular and paracellular mechanisms [31]. Mammalian cells lack specific cell surface transporters for mannitol, but passive diffusion across the cellular layer of the gut still occurs. Due to its small size and inert nature, mannitol can also be transported paracellularly through intact tight junctional pores in healthy tissue [32]. No doubt, mannitol passage can also occur non-specifically when there is overt damage to the intestinal wall. Measuring urinary mannitol excretion over a 24-hour period following ingestion of the probe (as done here) does not allow us to distinguish among these possibilities. Thus, the current study does not elucidate how Aquamin^®^ affects the permeability barrier in the gut.

To gain insight into Aquamin^®^’s mechanism(s) of action, our previous organoid culture studies and two recent interventional trials are instructive. In organoid culture, inclusion of Aquamin^®^ in the medium had only a modest effect on tight junctional protein expression, but strongly up-regulated adherens junction and desmosomal cadherins as well as other proteins necessary for cell-cell adhesion [16–21]. Basement membrane components and proteins that mediate cell-matrix adhesion via hemidesmosomes and focal adhesions were also up-regulated [16,17,19]. Finally, proteins that make up the mucinous layer were enhanced in the presence of Aquamin^®^ [16,17,20,21]. Not surprisingly, given this wide range of affected proteins, an increase in TEER as well as an increase in tissue cohesion was seen in Aquamin^®^-treated organoids [18–20]. Of interest, the response to Aquamin^®^ was as robust in organoids derived from healthy tissue as it was in UC-derived organoids [16,17]. These data suggest that, in organoid culture at least, minerals in the algae-derived product act on basic physiological processes that drive barrier protein generation. Enhanced barrier protein expression likely makes the tissue more resistant to inflammatory injury initially and allows the tissue to regain normal barrier function more rapidly when injury does occur. A role for Aquamin^®^ that is independent of a direct effect on inflammation is, therefore, suggested.

In the same organoid culture studies, a proteomic platform was used to assess a wide range of protein changes in response to Aquamin^®^ treatment. Several proteins with anti-inflammatory potential were induced while other proteins that are known to promote inflammation were downregulated [16,17,20,21]. Thus, while we hypothesize that the major effect of Aquamin^®^ is on barrier protein expression, a direct anti-inflammatory role is also possible. These two effects are not mutually exclusive. Either could lead to improved permeability control.

In the two interventional trials [22,23], daily ingestion of Aquamin^®^ for either 90 or 180 days altered many of the same barrier proteins and inflammation-related proteins that were affected in organoid culture. This supports the conclusions reached from the organoid culture studies. In addition, however, Aquamin^®^ ingestion was also shown to alter the microbial profile in the colon and reduce the level of several toxic bile acids [25]. Both effects could contribute to improved intestinal permeability and gut health. Thus, caution should be taken in interpreting Aquamin^®^’s primary effect as being simply due to increased barrier protein expression or a direct effect on inflammation. Ultimately, it is not unreasonable to suggest that a multi-mineral product could affect gastrointestinal health through multiple independent or overlapping mechanisms.

As part of the current study, as well as in our two previous trials [22,23], all subjects had a serum metabolic panel performed at the start and end of the interventional period. In none of the three trials were detectable changes observed in any of the metabolites that would indicate a safety concern. Of more interest, in both the recent trial with UC patients [23] and in the current study, serum ALP levels decreased over the course of treatment with Aquamin^®^. In the current study (following a 90-day intervention), the decline was statistically significant (p<0.05). In the recent trial with UC patients [23], the decline in serum ALP was seen after both 90 days and 180 days of intervention but it reached a level of statistical significance only after 180 days. In that study, a second assay measuring the intestine-specific isoform of ALP (ALPI) in serum demonstrated a comparable decline over the 180-day treatment period. In parallel with the decline in serum ALPI levels, colonic tissue ALPI levels were higher after 180 days of Aquamin^®^ treatment period than in pretreatment samples [23]. This was not observed in UC patients receiving placebo. While the significance of this finding is not fully understood, ALP is localized on the brush border of enterocytes, where it has several possible functions that could contribute to the anti-inflammatory shield in the gastrointestinal tract [33,34], or directly help to regulate permeability [34]. ALPI plays a crucial role in preserving gut barrier integrity by detoxifying microbial components and reducing inflammation. Studies indicate that upregulation or supplementation of ALPI can improve barrier dysfunction and support intestinal health [33,34]. In addition, elevated serum ALP levels are often seen in UC as well as in CD and it is often accompanied by primary sclerosing cholangitis (PSC) [35]. Whether Aquamin^®^-induced changes in ALP expression simply reflect improved colon tissue health or directly contribute to these improvements remains to be determined. Likewise, while serum ALP is a useful prognostic indicator in PSC [36], its pathophysiological role is unclear.

While the current study provides the first direct evidence that Aquamin^®^ can improve barrier function in humans, the study has limitations. First, as already noted, the use of mannitol alone as a probe for permeability status can identify alterations but does not address mechanism of action.

Importantly, it is not known if large moieties such as intact microbial cells, cellular components or food-derived allergens would show the same reduced paracellular passage in response to Aquamin^®^ as was seen here with the monosaccharide. Including larger sugar molecules such as lactulose could be beneficial in this regard as some studies have demonstrated that the lactulose/mannitol ratio provides data not obtainable with mannitol alone [1–3]. We tried this approach but could not obtain reliable lactulose data with existing ELISA kits. While mannitol alone has certain limitations, the consistency of the findings presented here, and the results from past studies with this probe [37–40] clearly attest to its utility.

A second limitation relates to the small sample size and the short duration of intervention in the present study. When this study was initiated, we did not know how much more effective a 180-day treatment period with Aquamin^®^ would be compared to 90 days of treatment. Even a duration of 180 days may not be optimal. In future research, we hope to use a full-year treatment period. The fact that we have observed no study agent-related serious adverse events, no adverse changes in the metabolic panel read-outs and no tolerability issues either here or in two previous trials [22,23,25], supports moving forward with a larger trial and longer interventional period. Additionally, increasing the amount of Aquamin^®^ could enhance efficacy. To begin addressing these limitations, we have initiated a new trial in UC patients with a J-pouch (following ileal pouch-anal anastomosis) to study effect of Aquamin^®^ in the prevention of pouchitis (inflammation of the intestinal pouch lining), a common downstream complication of this procedure.

In summary, our previous *ex vivo* and *in vivo* studies have provided compelling evidence for improved barrier function in the gastrointestinal tract with Aquamin^®^. Here we present direct evidence for improvement in gastrointestinal barrier function. While these investigations were conducted primarily to support the development of Aquamin^®^ as an ancillary treatment for UC, barrier dysfunction is recognized as a pathophysiological component of many other gastrointestinal disorders. In addition to UC and CD, these include malabsorptive conditions such as celiac disease [41], IBS [42], and diseases affecting the liver and pancreas [43]. Importantly, increased intestinal permeability has also been observed in various neurological disorders—including Alzheimer’s disease, Parkinson’s disease, amyotrophic lateral sclerosis, and anxiety/depression [2]. Finally, recent studies have suggested that gastrointestinal barrier disruption can serve as a predictor of incipient transplant rejection in allogenic tissue transplantation [44]. Given Aquamin^®^’s demonstrated safety profile and potential to enhance barrier integrity, it may offer therapeutic benefit as an adjuvant in any of these settings. Future studies are warranted to explore its application beyond UC, targeting these diverse disease populations.

## Data Availability

All relevant data are included in the manuscript and its Supporting Information files.

## Acknowledgments

The study team gratefully acknowledges the Michigan Institute for Clinical and Health Research (MICHR), the Michigan Clinical Research Unit (MCRU), the Research Pharmacy, and the Clinical Trials Support Office (CTSO) at the University of Michigan for their essential support of this trial. We extend special thanks to Mitch Seymour of the MICHR IND/IDE Investigator Assistance Program (MIAP) for guidance with the IND process, and to Sherece Bank of the Clinical Research Management (CRM) team for assistance with the REDCap (Research Electronic Data Capture) database. We also thank our study coordinators, Constantine Nolan and Almo Regazi, for their invaluable contributions. We are grateful to Marigot Ltd. (Cork, Ireland) for providing Aquamin^®^ capsules as a gift. Most importantly, we thank the study participants (healthy volunteers and patients with ulcerative colitis), without whom this study would not have been possible.

## Supplementary Tables

**Supplementary Table 1:**
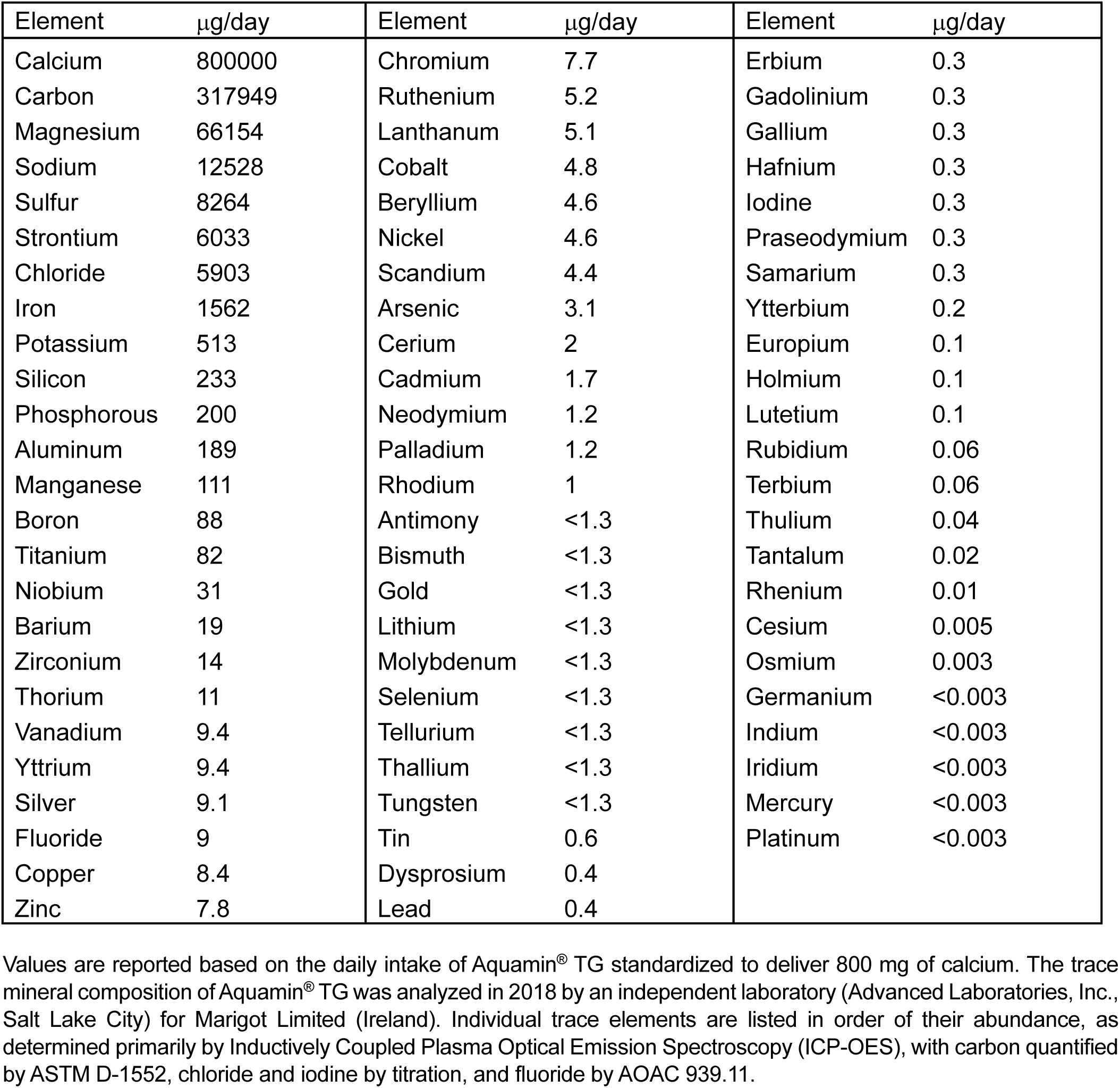
Elemental Composition and Daily Intake of Aquamin^®^ TG.

| Element | µg/day | Element | µg/day | Element | µg/day |
| --- | --- | --- | --- | --- | --- |
| Calcium | 800000 | Chromium | 7.7 | Erbium | 0.3 |
| Carbon | 317949 | Ruthenium | 5.2 | Gadolinium | 0.3 |
| Magnesium | 66154 | Lanthanum | 5.1 | Gallium | 0.3 |
| Sodium | 12528 | Cobalt | 4.8 | Hafnium | 0.3 |
| Sulfur | 8264 | Beryllium | 4.6 | Iodine | 0.3 |
| Strontium | 6033 | Nickel | 4.6 | Praseodymium | 0.3 |
| Chloride | 5903 | Scandium | 4.4 | Samarium | 0.3 |
| Iron | 1562 | Arsenic | 3.1 | Ytterbium | 0.2 |
| Potassium | 513 | Cerium | 2 | Europium | 0.1 |
| Silicon | 233 | Cadmium | 1.7 | Holmium | 0.1 |
| Phosphorous | 200 | Neodymium | 1.2 | Lutetium | 0.1 |
| Aluminum | 189 | Palladium | 1.2 | Rubidium | 0.06 |
| Manganese | 111 | Rhodium | 1 | Terbium | 0.06 |
| Boron | 88 | Antimony | <1.3 | Thulium | 0.04 |
| Titanium | 82 | Bismuth | <1.3 | Tantalum | 0.02 |
| Niobium | 31 | Gold | <1.3 | Rhenium | 0.01 |
| Barium | 19 | Lithium | <1.3 | Cesium | 0.005 |
| Zirconium | 14 | Molybdenum | <1.3 | Osmium | 0.003 |
| Thorium | 11 | Selenium | <1.3 | Germanium | <0.003 |
| Vanadium | 9.4 | Tellurium | <1.3 | Indium | <0.003 |
| Yttrium | 9.4 | Thallium | <1.3 | Iridium | <0.003 |
| Silver | 9.1 | Tungsten | <1.3 | Mercury | <0.003 |
| Fluoride | 9 | Tin | 0.6 | Platinum | <0.003 |
| Copper | 8.4 | Dysprosium | 0.4 |  |  |
| Zinc | 7.8 | Lead | 0.4 |  |  |

Values are reported based on the daily intake of Aquamin® TG standardized to deliver 800 mg of calcium. The trace mineral composition of Aquamin® TG was analyzed in 2018 by an independent laboratory (Advanced Laboratories, Inc., Salt Lake City) for Marigot Limited (Ireland). Individual trace elements are listed in order of their abundance, as determined primarily by Inductively Coupled Plasma Optical Emission Spectroscopy (ICP-OES), with carbon quantified by ASTM D-1552, chloride and iodine by titration, and fluoride by AOAC 939.11.

**Supplementary Table 2.** Health-related quality of life assessment (IBDQ) scoring.

| IBDQ Measures | Normal Range | Pre | Post |
| --- | --- | --- | --- |
| T-score (Total) | 32–224 | 191.6 ± 15.3 | 194.3 ± 20.5 |
| Bowel symptoms score | 10–70 | 61.4 ± 5.9 | 62.3 ± 7.2 |
| Systemic symptoms score | 5–35 | 27.5 ± 4.2 | 28.1 ± 4.7 |
| Emotional function score | 12–84 | 68.6 ± 7.0 | 69.8 ± 7.7 |
| Social function score | 5–35 | 34.1 ± 1.1 | 34.1 ± 2.1 |

Values represent Mean ± Standard Deviation for each respective measure; n = 8 subjects. The observed baseline IBDQ total score range was 172–200. A total IBDQ score of ≥ 170 is commonly used as a cutoff for clinical remission. The Inflammatory Bowel Disease Questionnaire (IBDQ) is a 32-item, patient-reported instrument assessing four domains: bowel symptoms, systemic symptoms, emotional function, and social function, using 7-point Likert-type scales. Higher scores indicate better quality of life; domain scores correspond to their specific item ranges.

**Supplementary Table 3.** Demographic Data for Subjects Enrolled in the 90-Day Study.

A. Study cohort demographics:
|  | Female |  | Male |  | Both Genders |  |
| --- | --- | --- | --- | --- | --- | --- |
| **Ethnic Category** | N | % | N | % | Total | % |
| Hispanic or Latino | 1 | 3.23% | 2 | 6.45% | 3 | 9.68% |
| Not Hispanic or Latino | 14 | 45.16% | 13 | 41.94% | 27 | 87.10% |
| Unknown | 0 | 0.00% | 0 | 0.00% | 0 | 0.00% |
| Prefer not to answer | 1 | 3.23% | 0 | 0.00% | 1 | 3.23% |
| **Total** | **16** | **51.61%** | **15** | **48.39%** | **31** | **100.00%** |
| **Racial Category**<br>(single category per participant) | N | % | N | % | Total | % |
| American Indian/Alaska Native | 1 | 3.23% | 0 | 0.00% | 1 | 3.23% |
| Asian | 2 | 6.45% | 1 | 3.23% | 3 | 9.68% |
| Native Hawaiian or Pacific Islander | 0 | 0.00% | 0 | 0.00% | 0 | 0.00% |
| Black or African American | 0 | 0.00% | 1 | 3.23% | 1 | 3.23% |
| White | 13 | 41.94% | 13 | 41.94% | 26 | 83.87% |
| Other | 0 | 0.00% | 0 | 0.00% | 0 | 0.00% |
| Unknown | 0 | 0.00% | 0 | 0.00% | 0 | 0.00% |
| **Total** | **16** | **51.61%** | **15** | **48.39%** | **31** | **100.00%** |
| **Age at Enrollment Category** | N | % | N | % | Total | % |
| 18 - 21 years | 4 | 12.90% | 1 | 3.23% | 5 | 16.13% |
| 22 - 29 years | 3 | 9.68% | 3 | 9.68% | 6 | 19.35% |
| 30 - 39 years | 3 | 9.68% | 5 | 16.13% | 8 | 25.81% |
| 40 - 49 years | 4 | 12.90% | 3 | 9.68% | 7 | 22.58% |
| 50 - 59 years | 1 | 3.23% | 2 | 6.45% | 3 | 9.68% |
| 60 - 69 years | 1 | 3.23% | 1 | 3.23% | 2 | 6.45% |
| 70 - 79 years | 0 | 0.00% | 0 | 0.00% | 0 | 0.00% |
| > 80 years | 0 | 0.00% | 0 | 0.00% | 0 | 0.00% |
| **Total** | **16** | **51.61%** | **15** | **48.39%** | **31** | **100.00%** |

B. Gender and Age:
|  | <u>Gender</u> | <u>Age (Y)</u> |
| --- | --- | --- |
| Healthy (n=19) | M:9 / F:10 | 31.7±12.0 |
| UC (n=12) | M:6 / F:6 | 41.9±12.7 |

C. Body Mass Index:
|  | <u>BMI - Weight/Height<sup>2</sup> (Kg/m<sup>2</sup>)</u> |  |
| --- | --- | --- |
|  | <u>Pre</u> | <u>Post</u> |
| Healthy (n=12) | 24.3±4.0 | 24.6±4.4 |
| UC (n=8) | 26.5±5.7 | 27.6±5.3 |

**Supplementary Table 4.** Summary of Average Mineral Intake and Recommended Levels.

| **Nutrient (Unit)** | **Healthy Subjects** |  |  |  | **Subjects with UC** |  |  |  |
| --- | --- | --- | --- | --- | --- | --- | --- | --- |
|  | Eaten | Suggested Target | Min | Max | Eaten | Suggested Target | Min | Max |
| **Calcium (mg)** | 1089.0 ± 526.4 | 1120.0 ± 164.3 | 384 | 1775 | 1169.0 ± 415.1 | 1028.6 ± 75.6 | 565 | 1811 |
| **Copper (mg)** | 1.3 ± 0.6 | 0.9 ± 0.0 | 0.7 | 2 | 1.5 ± 0.4 | 0.9 ± 0.0 | 0.7 | 2 |
| **Iron (mg)** | 11.8 ± 6.0 | 12.8 ± 4.5 | 5.7 | 20 | 13.6 ± 3.2 | 13.7 ± 5.3 | 8.3 | 18 |
| **Magnesium (mg)** | 293.0 ± 110.9 | 372.0 ± 39.0 | 145 | 420 | 377.1 ± 119.6 | 345.7 ± 44.3 | 191 | 530 |
| **Phosphorus (mg)** | 1173.4 ± 475.8 | 920.0 ± 301.2 | 597 | 1890 | 1436.6 ± 417.0 | 700.0 ± 0.0 | 705 | 1918 |
| **Potassium (mg)** | 2325.4 ± 1126.2 | 4700.0 ± 0.0 | 1195 | 3916 | 2701.9 ± 686.1 | 4700.0 ± 0.0 | 1773 | 3796 |
| **Selenium (mcg)** | 103.0 ± 51.2 | 55.0 ± 0.0 | 48 | 187 | 79.7 ± 26.0 | 55.0 ± 0.0 | 53 | 128 |
| **Zinc (mg)** | 9.3 ± 3.6 | 9.6 ± 1.3 | 5.4 | 15 | 11.2 ± 2.9 | 8.9 ± 1.5 | 5.5 | 14 |
| **Vitamin D (mcg)** | 2.2 ± 0.7 | 15.0 ± 0.0 | 1.6 | 3 | 5.8 ± 3.8 | 15.0 ± 0.0 | 1.9 | 12 |

Participants (n = 5 for healthy subjects and n = 7 for UC subjects) reported their food and beverage intake, including quantities and frequency, over the past year using the NIH Diet History Questionnaire III (DHQ3), developed by the National Cancer Institute (NCI). These responses were used to calculate average daily intakes of nutrients and food groups. Average intakes were then compared to current dietary guidelines based on age and sex, as outlined in the Dietary Guidelines for Americans<sup>1</sup>. The table presents a subset of mineral intakes calculated using the DHQ3 database<sup>a</sup>, expressed as means and standard deviations. These are the only minerals detected by DHQ3.
Note: Vitamin D intake in DHQ3 is calculated from food, beverage, and supplement data only. Sun exposure and cutaneous vitamin D synthesis are not assessed by DHQ3.
<sup>1</sup>Dietary Guidelines for Americans. https://www.dietaryguidelines.gov/resources/2020-2025-dietary-guidelines-online-materials
<sup>a</sup>Mineral values were calculated using the Diet History Questionnaire version 3 (DHQ3) database.

**Supplementary Table 5.**
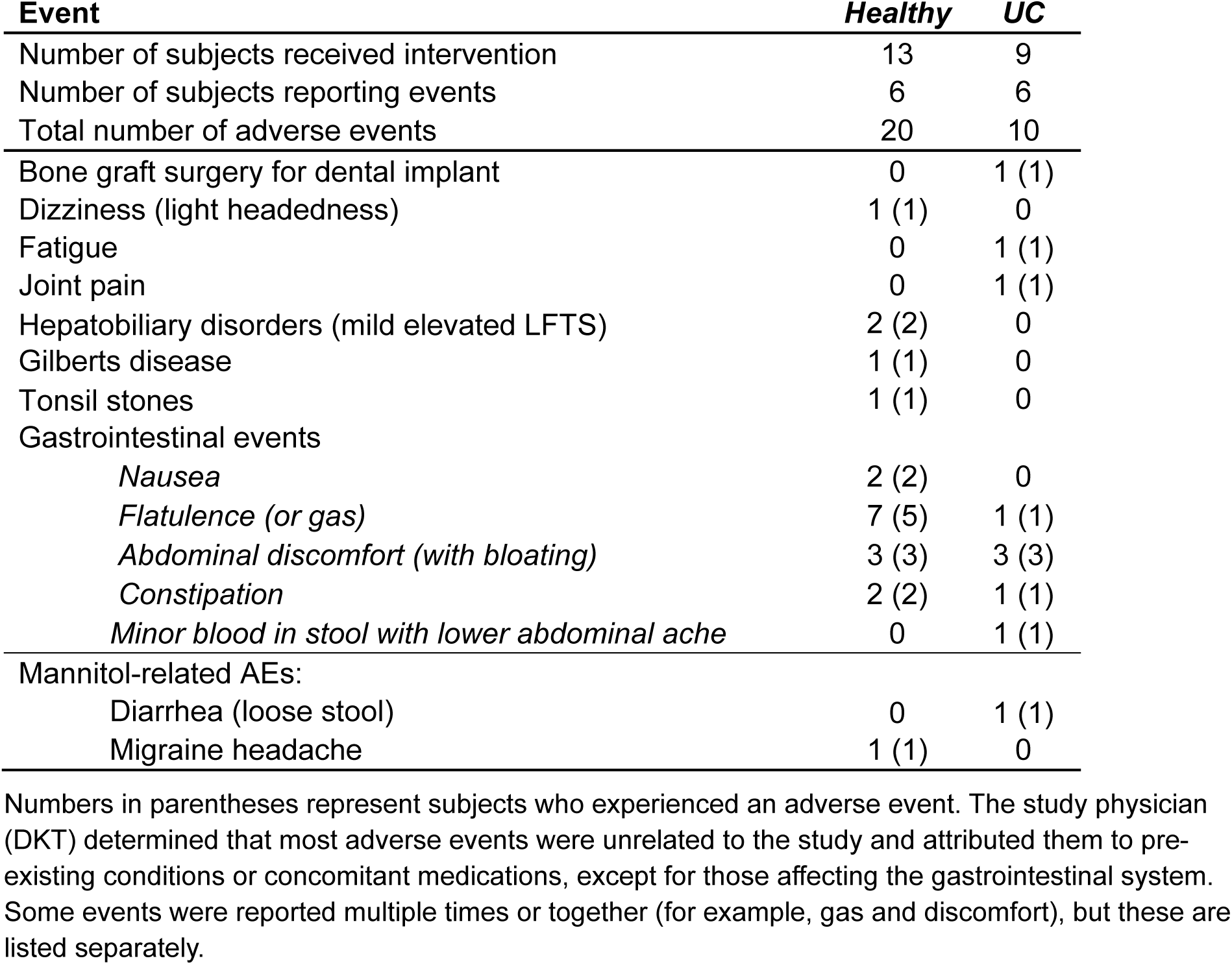
Treatment-Related Reportable Adverse Events.

